# Short-term change in air pollution following the COVID-19 state of emergency: A national analysis for the United States

**DOI:** 10.1101/2020.08.04.20168237

**Authors:** Pooja Tyagi, Danielle Braun, Benjamin Sabath, Lucas Henneman, Francesca Dominici

## Abstract

Lockdown measures taken in response to the COVID-19 pandemic produced sudden social and economic changes. We examined the extent of air pollution reduction that was attained under these extreme circumstances, whether these reductions occurred everywhere in the US, and the local factors that drove them. Employing counterfactual time series analysis based on seasonal autoregressive integrated moving average models, we found that these extreme lockdown measures led to a reduction in the weekly PM_2.5_ average by up to 3.4 µg m^-3^ and the weekly NO_2_ average by up to 11 ppb. These values represent a substantial fraction of the annual mean NAAQS values of 12 µg m^-3^ and 53 ppb, respectively. We found evidence of a statistically significant decline in NO_2_ concentrations following the state-level emergency declaration in almost all states. However, statistically significant declines in PM_2.5_ occurred mostly in the West Coast and the Northeast. Certain states experienced a decline in NO_2_ but an increase in PM_2.5_ concentrations, indicating that these two pollutants arise from dissimilar sources in these states. Finally, we found evidence that states with a higher percentage of mobile source emissions prior to the emergency measures experienced a greater decline in NO_2_ levels during the pandemic. Although the current social and economic restrictions are not sustainable, our results provide a benchmark to estimate the extent to which air pollution reductions can be achieved. We also identify factors that contributed to the magnitude of pollutant reductions, which can help guide future state-level policies to sustainably reduce air pollution.

**Significance statement:** We quantified the reduction in air pollution levels achieved under the extreme social and economic measures that were put into place as part of COVID-19 state-level emergency declarations. We found a reduction in the weekly average PM_2.5_ of up to 3.4 µg m^-3^ and the weekly average of NO_2_ of up to 11 ppb. These values represent a substantial fraction of the annual mean NAAQS values of 12 µg m^-3^ and 53 ppb, respectively. States with a larger fraction of mobile source emissions (e.g., air and road traffic) prior to the pandemic experienced larger declines in NO_2_ emissions, whereas PM_2.5_ decline was seen in areas with a higher pre-pandemic proportion of emissions from mobile, stationary (e.g., industrial) and fire sources.

## Introduction

There is consistent evidence that short- and long-term exposure to fine particulate matter (PM_2.5_) and nitrogen dioxide (NO_2_) increases the risk of mortality, hospitalization, and other adverse health outcomes (1–6). Furthermore, several studies have provided preliminary evidence that long-term air pollution exposure increases the risk of hospitalization and death among individuals with COVID-19 (4, 7). Yet, despite the large body of evidence that indicates the current National Ambient Air Quality Standards (NAAQS) for PM_2.5_ may not sufficiently protect public health (8–11), the US Environmental Protection Agency (EPA) announced a sweeping relaxation of environmental rules in response to the COVID-19 pandemic.

The United States mitigates air pollution through a combination of federal, state, and local air pollution regulations (12). For example, the federal government sets emissions standards and the NAAQS. They also require states to prepare State Implementation Plans (SIPs) that detail emissions reductions strategies for areas that are not in compliance with the NAAQS (non-attainment areas). SIPs use air quality models to demonstrate how regulating local emissions sources helps a non-attainment area meet the NAAQS. The patchwork of regulations, emission sources, geographical variability, and meteorology, results in varying air pollution concentrations by location.

Several studies have examined the impact of a sudden intervention on changes in air pollution (see (13) for a review). For example, researchers used interrupted time-series designs to quantify the impact of the 1990 Dublin coal ban (14) and regression discontinuity to identify the arbitrary spatial impact of the China Huai River Policy (15). An important feature of these studies is that they investigated abrupt and localized changes across a relatively short time span (Dublin coal ban) and spatial scale (Huai River policy) (16). Because of the abrupt nature of these interventions, defining a hypothetical experiment in these studies was straightforward. For example, in the Dublin study, the institution of the ban represented the intervention condition, with the control condition being no ban, and the effect of interest was that of instituting the ban versus not instituting the ban.

Similarly, we examined the effect of the abrupt lockdown measures implemented in response to the COVID-19 pandemic, which produced sudden and significant changes in how society functions, with decreases in road traffic, air traffic, and economic activity (17). This provided us with an unprecedented opportunity to implement a quasi-experimental design with a well-defined control condition (no pandemic) to estimate the changes in air pollution as a result of the implementation of these extreme measures (see (18) for a discussion of strengths and limitations of a quasi-experimental design). Furthermore, the spatial heterogeneity in the extent to which air pollution levels changed as a result of the lockdown measures allowed us to identify factors contributing to these changes.

In this study, we had two main scientific objectives: 1) use state-of-the-art time series approaches for counterfactual forecasting to estimate the weekly state-level deviations between counterfactual (e.g., absent the pandemic) and observed levels of PM_2.5_ and NO_2_ from January 1, 2020 to April 23, 2020; and 2) investigate which state-level characteristics, including emissions sources, contributed the most to these changes, while adjusting for geography and population density. We decided to estimate counterfactual values starting in January 2020 to allow for the identification of the time point when these counterfactual predictions began deviating from observed concentrations. We then compared the onset of this deviation to the timing of different interventions, including declaration of a state of emergency, non-essential business closures, and shelter-in-place orders (see Figures S8 and S9 in the supplemental material). We chose the date of the state-level emergency declarations as the primary intervention for this analysis because it was the earliest intervention that most closely aligned with the decline in air pollution levels. We used April 23, 2020 as the end date for our study because all state-level emergency declarations occurred at some point during this period, and the time range gave us more than four weeks of post-intervention follow-up time for each state.

## Results

### Short-term change in air pollution following the COVID-19 state of emergency

To estimate the changes in air pollution levels (PM_2.5_ and NO_2_) affected by the COVID-19 state-level emergency declaration (denoted by Δ), we implemented Seasonal Autoregressive Integrated Moving Average (SARIMA) models for multivariate time series data (see Materials and Methods for details). More specifically, we trained SARIMA models on historical state-level data (January 1, 2015 to December 31, 2019) to forecast the counterfactual (i.e., pandemic did not occur) weekly pollutant concentrations for a 16-week period (January 1, 2020 to April 23, 2020). For each state, we estimated the weekly deviations of the observed pollutant concentrations from their counterfactual values (and associated statistical uncertainty using bootstrapping) for the same 16-week period (i.e., weekly observed values minus weekly predicted values for 16 weeks). Then, we estimated Δ by taking the difference Δ*_before_*−Δ*_after_*, where Δ*_before_* is the median of these weekly deviations *before* the date of the declaration of the state emergency (denoted by T) and Δ*_after_* is the median of these weekly deviations *after* T. A negative estimated value of Δ indicates that air pollution levels declined as a result of the state-level emergency (e.g., the deviations are closer to zero before the intervention, but after the intervention, the weekly predictive values assuming that the pandemic did not occur are higher than the observed ones). We assessed statistical uncertainty using bootstrap. This analysis was performed separately for PM_2.5_ and NO_2_ in each state.

For most states, the differences between the SARIMA counterfactual predictions (i.e., assuming the pandemic did not occur) and the observed pollutant values were close to zero during the period before the state-level emergency declaration (Figures 1-2), but there were significant deviations following the intervention lockdown measures.

**Figure 1.**
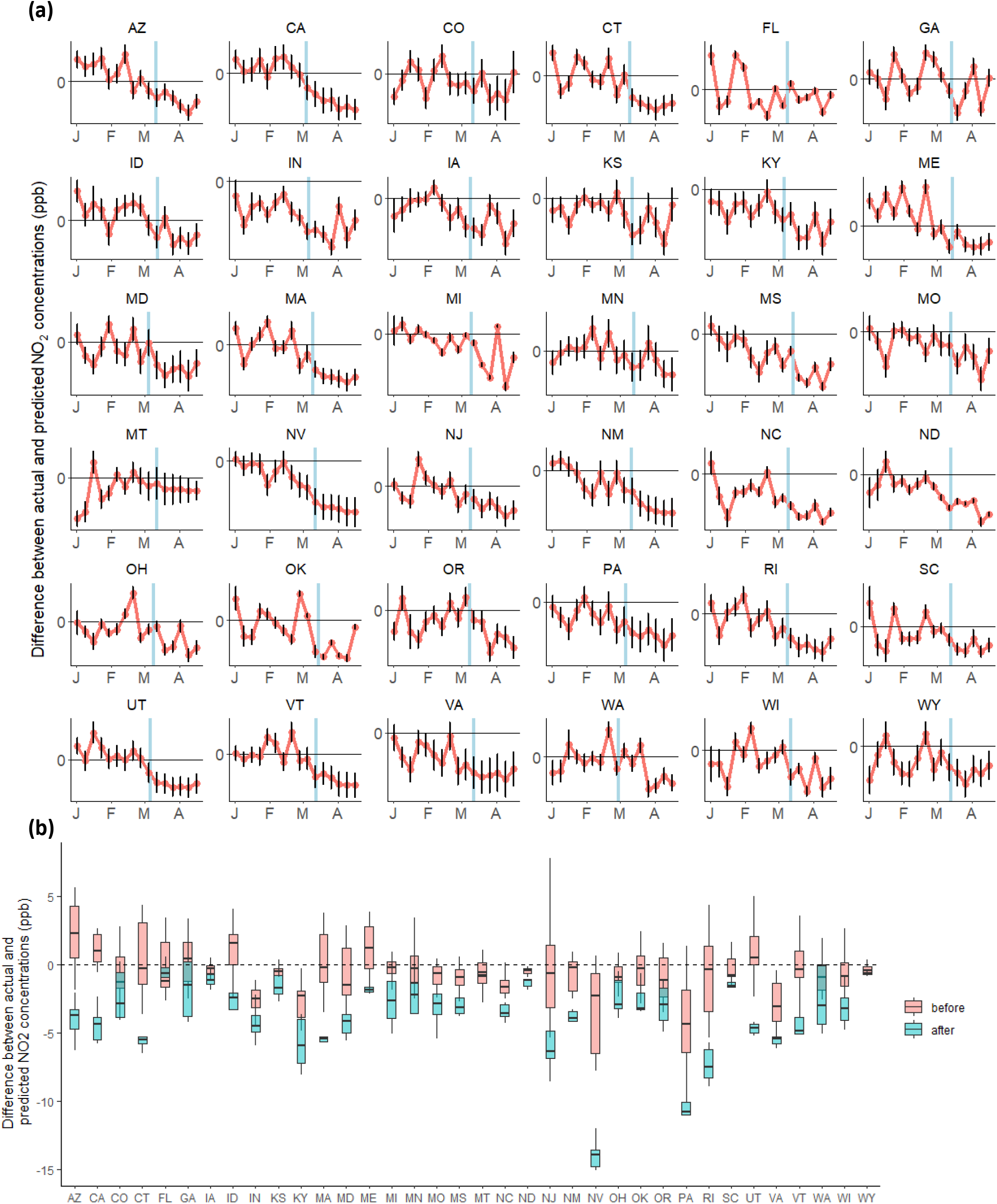
(a) Weekly deviations between observed NO_2_ concentrations and counterfactual predictions (e.g., absent the pandemic) for each state. The counterfactual predictions were made for 16 weeks from January 1 to April 23, 2020. The blue vertical line marks the date of the declaration of a state of emergency in each state. (b) Boxplots of the weekly deviations for the weeks before (pink) and for the weeks after (blue) the date of the declaration of a state of emergency in each state.

**Figure 2.**
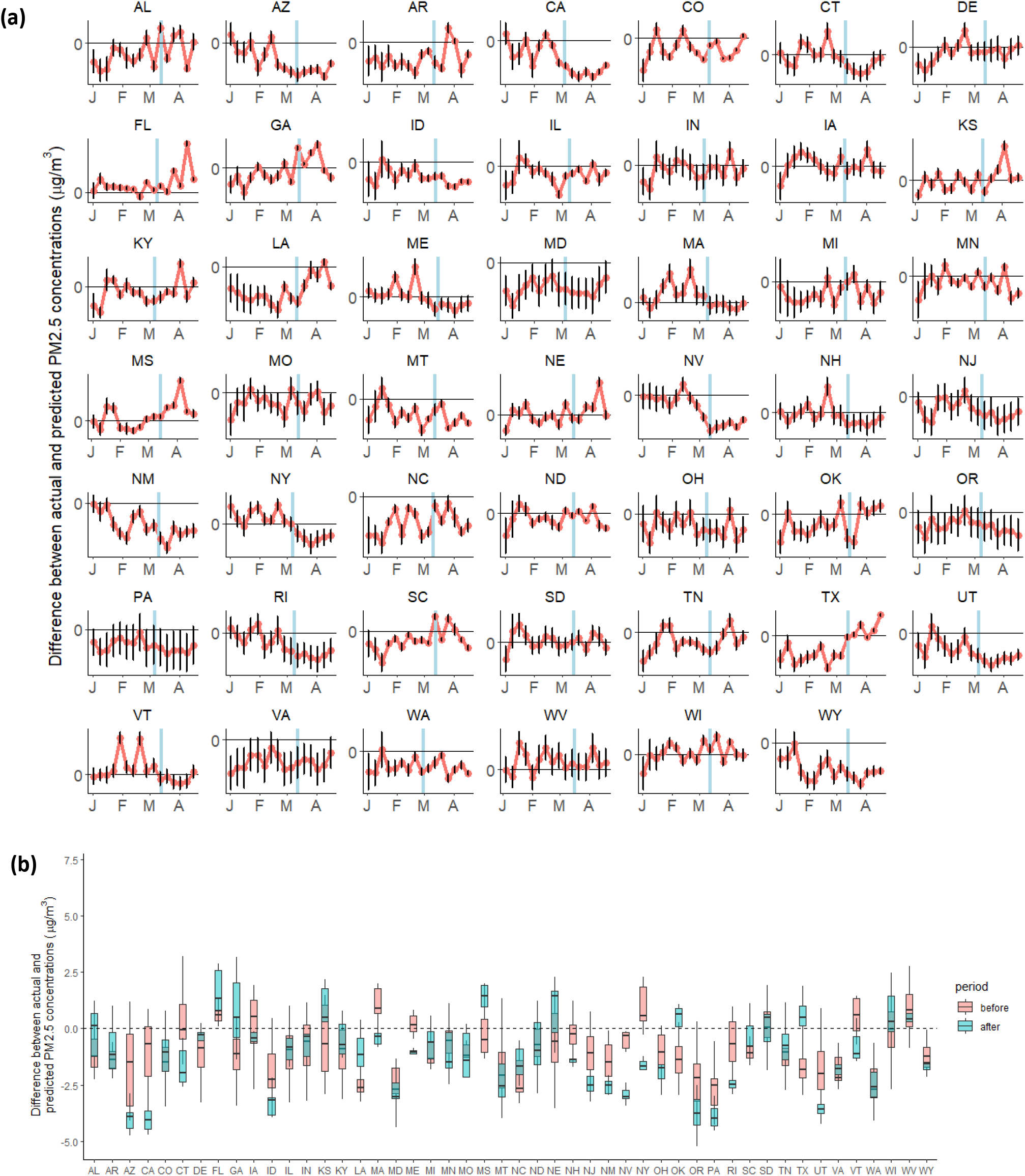
(a) Weekly deviations between observed PM_2.5_ concentrations and counterfactual predictions (e.g., absent the pandemic) for each state. The predictions were made for 16 weeks from January 1 to April 23, 2020. The blue vertical line marks the date of the declaration of a state of emergency in each state. (b) Boxplots of the weekly deviations for the weeks before (pink) and for the weeks after (blue) the date of the declaration of a state of emergency in each state.

We found evidence of a statistically significant decrease in NO_2_ concentrations following the declaration of a state of emergency in 34 of the 36 states that were investigated (Figure 1 and Table S3). However, we found evidence of a statistically significant decline in PM_2.5_ concentrations in only 16 of the 48 states that were studied, including New York and other states in the Northeast and West Coast (Figures 2-3 and Figure S7).

**Figure 3.**
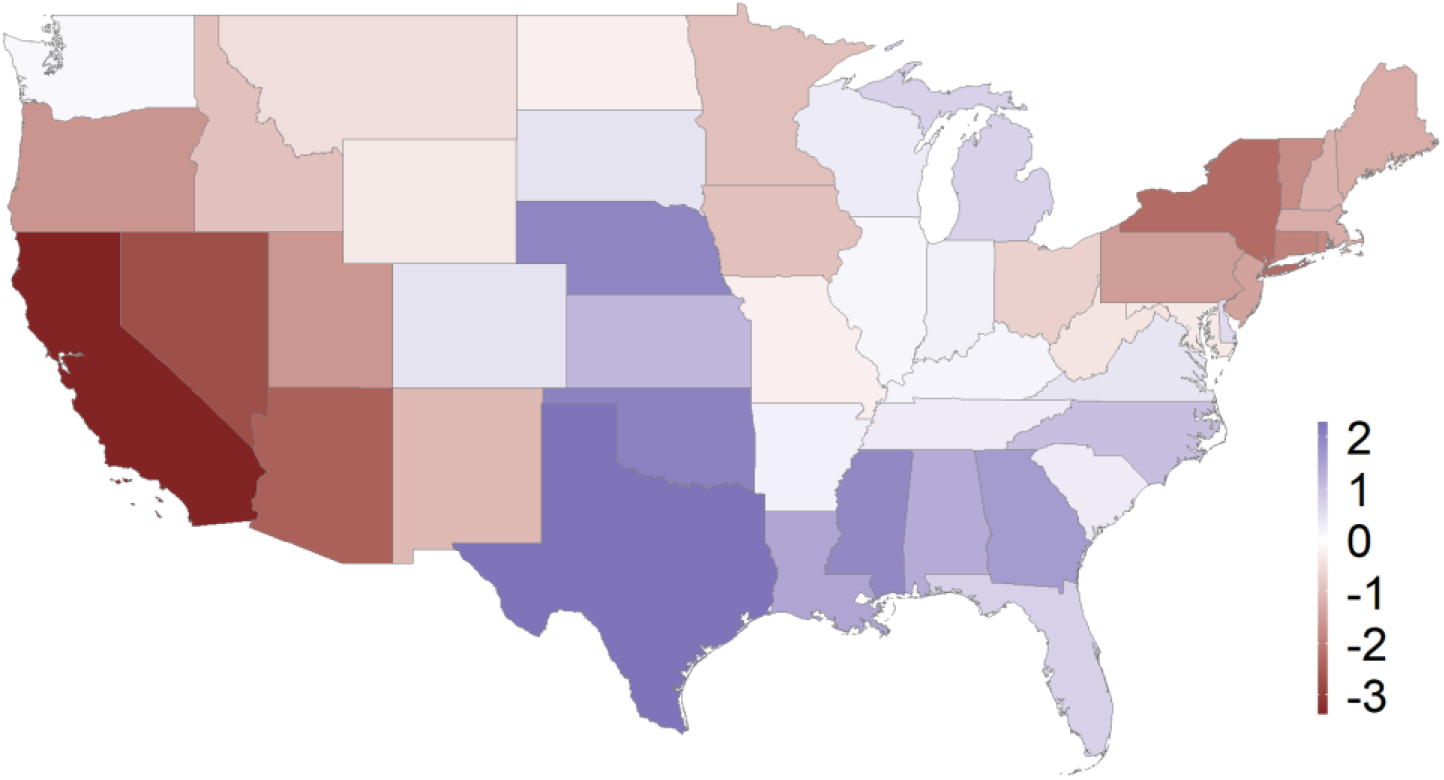
Median change in in PM_2.5_ following the state-level emergency declaration for each state (Δ). A negative estimated value of Δ indicates that air pollution levels declined as a result of the state-level emergency declaration.

Figure 4 shows ρ, defined as the ratio of the estimated Δ for NO_2_ divided by the estimated Δ for PM_2.5_. This ratio reflects the discrepancy in the magnitude and direction of change in the concentrations of NO_2_ and PM_2.5_ following state emergency declaration. More than one-third of the states had ρ < 0, i.e., these states experienced a decrease in NO_2_ and a simultaneous increase in PM_2.5_. The contrast between the pattern of change of NO_2_ and PM_2.5_ following state-level emergency declarations suggests that dominating sources of these two pollutants are different in those states.

**Figure 4.**
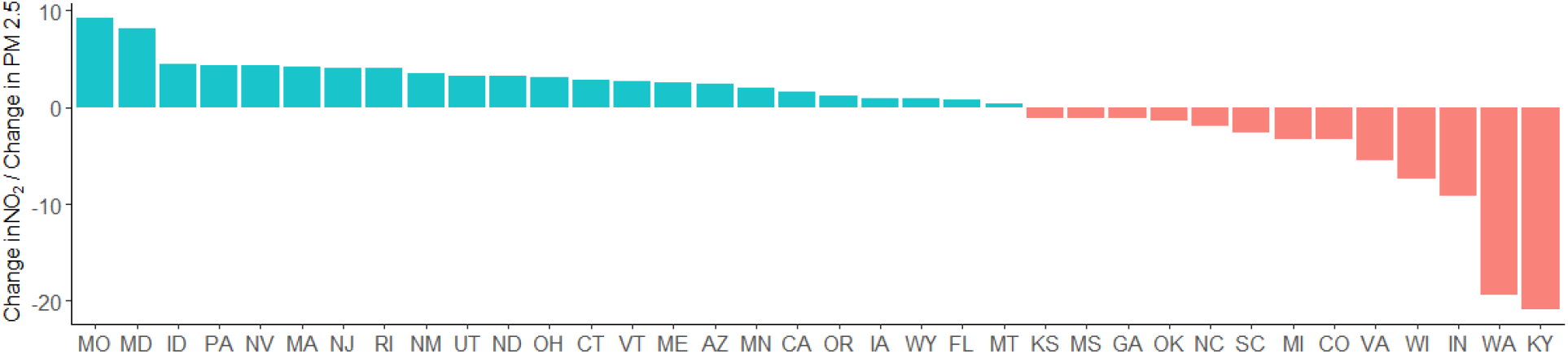
Ratio of the estimated Δ for NO_2_ divided by the estimated Δ for PM_2.5_ (ρ). A negative ratio implies that the change in NO_2_ following the declaration of the state of emergency was in the opposite direction of the corresponding changes for PM_2.5_ (i.e., one pollutant increased while the other decreased). For example, in KY, we found a decline in NO_2_ but an increase in PM_2.5_ following the state-level emergency declaration

### State-level factors may explain the heterogeneity in air pollution declines across states

To ascertain which state-level factors might explain the heterogeneity in the extent to which the air pollution declined across states, we fit weighted regression models with the estimated Δ as the dependent variable, and geography, population density and sources of emission as predictors. Details on the regression model can be found in the Materials and Methods. We found that the proportions of annual emissions from a state’s stationary (e.g., industrial processes), mobile (e.g., road and air traffic), and fire sources (e.g., agricultural field burning) have statistically significant and positive associations with the change in PM_2.5_ concentration (Table 1). In contrast, only the proportion of annual emissions from state mobile sources was a statistically significant predictor of the change in NO_2_ concentration (Table 1). These results indicate that states with a higher fraction of annual emissions from mobile sources experienced a greater decline in NO_2_ concentrations. For PM_2.5_, the higher the fraction of emissions from any of the three sources (stationary, mobile, or fire), the greater the decline in PM_2.5_.

**Table 1.** Estimated coefficients, 95% confidence intervals (CI) and P-values for variables included in the weighted linear regression analysis. The categorical variable *Region* refers to the geographical regions of the United States and has four categories as defined by the US Census, *Region 1: Northeast*, *Region 2: Midwest*, *Region 3: South* and *Region 4: West*, and the corresponding coefficients are reported using *Region 1* as reference.

|  | NO<sub>2</sub> change |  |  | PM<sub>2.5</sub> change |  |  |
| --- | --- | --- | --- | --- | --- | --- |
| | $\beta$ | 95% CI | P-value | $\beta$ | 95% CI | P-value |
| Fire | 1.089 | (-0.903,3.082) | 0.27 | 1.065 | (0.030, 2.1) | 0.04 |
| Mobile | 1.655 | (0.131, 3.178) | 0.03 | 1.240 | (0.29, 2.19) | 0.01 |
| Stationary | 1.063 | (-0.469,2.596) | 0.17 | 0.886 | (0.012, 1.760) | 0.05 |
| Population density | 0.000 | (-0.003,0.002) | 0.72 | -0.001 | (-0.002,.0003) | 0.18 |
| Region 2 | -2.883 | (-4.485, -1.282) | 0.00 | -1.147 | (-1.933, -0.360) | 0.01 |
| Region 3 | -3.310 | (-5.198, -1.423) | 0.00 | -2.248 | (-2.907, -1.589) | 0.00 |
| Region 4 | -0.748 | (-3.212, 1.716) | 0.54 | 0.095 | (-0.913, 1.103) | 0.85 |

## Discussion

In this study, we found that, following the declaration of a state of emergency, NO_2_ concentrations showed a statistically significant decline in 34 of the 36 states included in this analysis. In contrast, PM_2.5_ concentrations declined in only 16 of 48 states included in this analysis. These 16 states are located in the Northeast and on the West Coast. Furthermore, we found that the proportion of a state’s annual emissions from mobile sources is a statistically significant factor in NO_2_ changes in response to the state emergency declaration. For PM_2.5_ reductions, all three sources—mobile, stationary, and fire—were statistically significant predictors in driving this change. We concluded that state of emergency declarations implemented in response to the COVID-19 pandemic predominantly affected mobile sources (e.g., cancelled flights and reduced traffic) (19) and led to a decline in NO_2_. However, because the major sources of PM_2.5_ are stationary (e.g., industrial fuel combustion), these were less affected by state-level emergency declarations (Table S2).

SARIMA models have some advantageous features compared to other statistical approaches. Recent studies have used t-tests (20), a robust difference approach (21), and linear regression (22) to study the changes in US air pollution attributable to the COVID-19 shutdown. Bekbulat et al. used temporal correction in their robust differences approach (21) (not peer-reviewed on August 03, 2020) and Venter et al. included meteorological factors in their regression (22). The latter was a global study of air pollution changes during the pandemic, which found that a decline in NO_2_ in the United States occurred on a national level. However, these methods do not directly incorporate the correlations between observed pollutant concentrations, and trends and seasonality in the data. We accounted for both factors using SARIMA models. Any contribution to the data from generally decreasing air pollution trends and weather seasonality must be removed to best estimate the effect of pandemic-related extreme measures on air pollution. By further combining SARIMA with bootstrapping, we were able to quantify the uncertainty in the estimated mean predictions without any parametric assumptions.

We note that our counterfactual predictions of pollutant concentrations assume that the trend and seasonality during the last five years (i.e., the training period for the model) persisted during the prediction period (January 1, 2020 to April 23, 2020). Another assumption was that the relationship between meteorological variables used in the SARIMA model (temperature, humidity, and precipitation) and the pollutant concentrations were the same in both the training and prediction periods (23, 24).

In contrast to other studies, we did not *a priori* divide our data into pre- and post‒COVID-19 periods. We used January 1, 2015 to December 31, 2019 as historical data and then used the SARIMA model to predict the counterfactual pollutant levels during the 16-week period from January 1, 2020 to April 23, 2020, under the hypothesis that neither the pandemic nor the state emergency declaration occurred. In other words, first we estimated the changes between observed and counterfactual air pollution levels for the whole study period of 16 weeks. We then looked *a posteriori* to determine if the NO_2_ or PM_2.5_ declines coincided with state-level emergency declarations.

We also explored other measures, and found that, in most states, the declines in NO_2_ or PM_2.5_ occurred even before shelter-in-place orders and non-essential business closures (Figures S8 and S9). This highlights that economic activity and mobility were reduced even before these state measures were implemented (17).

By estimating the maximum decline in the median pollutant concentrations following state-level emergency declaration, we found that the extreme measures taken during the pandemic decreased the weekly PM_2.5_ average by up to 3.4 µg m^-3^ and the weekly NO_2_ average by up to 11 ppb. These values represent a substantial fraction of the annual mean NAAQS values of 12 µg m^-3^ and 53 ppb, respectively. This suggests the significant potential to reduce NO_2_ concentrations by reducing mobile NO_2_ emissions, and PM_2.5_ concentrations by reducing mobile sources, stationary sources, and fire-related emissions. For comparison, a recent study investigated the effect of lockdown in urban China, using difference-in-difference approach; they found a decline of 14 µg m^-3^ in locked-down cities compared to cities that did not implement a lockdown (25). The cities in that study had baseline PM_2.5_ concentrations four times higher than the safe limits set by the World Health Organization, which may have been partly responsible for a larger decline after lockdown compared to what we observed in the United States.

Our study results support the effectiveness of state-level actions to reduce ambient levels of PM_2.5_ and NO_2_, and specifically, that restrictions on mobile sources of air pollution could decrease NO_2_ emissions even further in states where mobile sources constitute a larger proportion of annual emissions. In contrast, PM_2.5_ concentration reduction may not be as easily achieved through mobile source emission restrictions alone, especially in states where stationary sources contribute more to air pollution. In states where changes in PM_2.5_ and NO_2_ exhibited opposite trends (one increased while the other decreased), lowering the emission of NO_2_, by decreasing mobile source emissions for example, may not necessarily decrease PM_2.5_ concentrations.

**Study Limitations**. We relied on state-level concentration averages and the 2014 emissions inventory. While our study would benefit greatly from a more recent emissions inventory (or fine spatial emissions estimates during the interventions), to our knowledge, such data is not currently available publicly. Trading finer spatial resolution in the monitoring data—not averaging to the state level—may reveal important sub-state variability in lockdown impacts. Our approach also does not consider the spatial correlations between pollutant concentrations, which may help explain concentration changes in non-local pollutants such as PM_2.5_. Finally, data were available for 36 states for NO_2_ and 48 states for PM_2.5_, which limited the number of observations in the weighted regression model.

## Materials and Methods

### Data Acquisition

We gathered and harmonized data from several databases (Table S1). We obtained historical daily monitor data of PM_2.5_ and NO_2_ concentrations for January 1, 2015 to August 31, 2019 from the US EPA Air Quality System (26). We obtained current levels of these air pollutants for August 31, 2019 to April 23, 2020 from the EPA AirNow application programming interface (27). We aggregated weekly monitor data within each state. These data were available for 48 states for PM_2.5_ and 36 states for NO_2_. We obtained daily temperature, humidity, and precipitation data from the University of Idaho’s GRIDMET project, which were then aggregated to the state level using Google Earth Engine (28).

We obtained state-level source emissions totals from the National Emissions Inventory (29), and gathered information on population density and geographic region classification of the states from the United States Census Bureau (30, 31). Finally, we accessed the COVID-19 US State Policy Database (32) to extract information regarding the date of the COVID-19 state-level declaration of emergency, shelter-in-place orders and non-essential business closures for each state.

### Statistical Methods

#### Counterfactual forecasting of air pollution levels starting January 1, 2020

We conducted the following analyses separately for PM_2.5_ and NO_2_ for each state.

1. We fitted SARIMA models (33–35) to the historical data on weekly state-level air pollution levels (from January 1, 2015 to December 31, 2019).
2. We predicted the air pollution counterfactual levels (absent the pandemic) during a 16-week period from January 01, 2020 to April 23, 2020, and then took the average across bootstrap iteration (see point 4 below): 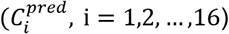
3. We estimated the weekly difference between the predicted and the observed values for this 16-week period: 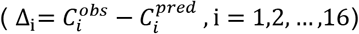
4. We quantified the statistical uncertainty of these estimated differences by bootstrap aggregating of SARIMA results (“bagged SARIMA”).

SARIMA models are autoregressive models often used to forecast time series where future observations are correlated with past observations (34, 35). They have the advantage of accounting for the trend and seasonality of the data. We also accounted for the effect of weather by including temperature, precipitation, and humidity as covariates in the SARIMA models. The basis of the SARIMA model is a linear regression of a response variable Y at time t against the past values (Y_t-1_, Y_t-2_, ….) of Y and the past forecast errors (ɛ_t_, ɛ_t-1_, …). A detailed example of this analysis for NO_2_ in California is provided in the supplementary materials. For each state, we created 1,000 time series bootstraps using Box-Cox and Loess-based decomposition (33). As shown in the supplementary materials, 1,000 bootstraps captured the original time series well (Figure S1) and allowed for reasonable computation time. We then fitted a new SARIMA model to each bootstrapped time series, providing new counterfactual predictions for the 16-week period. We averaged these predictions over the 1,000 bootstraps to generate a mean counterfactual prediction and confidence intervals for expected pollutant concentrations for each week (absent the pandemic). Finally, separately for each state, we calculated weekly deviations between the observed levels and the average of the counterfactual predictions (details in supplementary materials), as defined below.

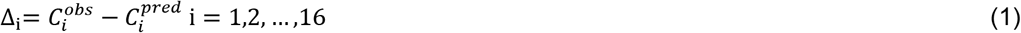

where, 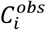 and 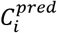 are the observed and the average of the counterfactual predictions for week *i* respectively. We used the R package auto.arima to select model coefficients with the best predictive capability based on bias-corrected Akaike Information Criterion (AIC) (36, 37) and then used the mean absolute scaled error (MASE) to evaluate the fit of the model.

We decided to start the counterfactual forecasting in January 2020 to allow for the identification of the time point when these counterfactual predictions began deviating from observed concentrations. We then compared the onset of this deviation to the date of COVID-19 declaration of a state of emergency for each state. We took this approach because fitting the SARIMA model using data until the date of a specific state intervention and predicting for the period following the state intervention would not allow for the assessment of whether the discrepancy between predicted and observed air pollution concentrations was attributable to the state intervention or the model’s poor predictive capability.

#### Model evaluation

In the supplementary material, we summarize information regarding the model evaluation. We found that MASE for the fitted models was less than 1 for each pollutant in each state, indicating that the SARIMA model outperformed one-step naïve forecasts (Figure S6).

#### Regression modeling to identify state-level factors contributing to heterogeneity in the air pollution declines across states

For each state and for each of the two pollutants (PM_2.5_ and NO_2_), we estimated the parameter Δ denoting the change in pollutant concentrations following the state intervention by calculating:

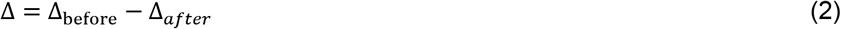

Where Δ*_before_* and Δ*_after_* denote the median of deviations between the weekly observed and counterfactuals before and after the date of the declaration of the state of emergency obtained from equation 1. We then fitted a weighted linear regression model with the estimated Δ as the outcome. We used the following independent variables: the proportion of emissions from fire sources, stationary sources, and mobile sources; population density; and region of the state.

#### Metric for comparison of pattern of change in NO_2_ and PM_2.5_

To identify the states with the most pronounced discrepancy between the pattern of change in PM_2.5_ and NO_2_, we calculated the ratio (ρ), defined as:

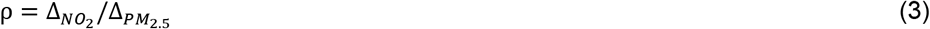

If ρ < 0, it implies that the two pollutants changed in opposite directions (i.e., one increased while the other decreased), and the larger the magnitude of ρ, the larger the discrepancy between the pollutants’ patterns of change.

## Data Availability

All data is publicly available from the sources presented in Table S1 in the supplementary information.

---

Supplementary Information

**Table S1.** Publicly available data sources used in the analysis.

| Data | Source |
| --- | --- |
| Daily monitor data for PM<sub>2.5</sub> and NO<sub>2</sub> emissions from January 1, 2015 to August 31, 2019 | US Environmental Protection Agency Air Quality System (26) |
| County-level daily data for PM<sub>2.5</sub> and NO<sub>2</sub> emissions after August 31, 2019 | EPA AirNow (27) |
| Daily temperature, humidity, and precipitation for all states | University of Idaho's GRIDMET project (28) aggregated to county from 4km x 4km rasters |
| State-level source emissions totals | National emissions inventory (29) |
| State population density and regions | 2010 Census data (30, 31) |
| Timing of state interventions | COVID-19 US state policy database (32) |

## SARIMA analysis for NO_2_ concentrations in California

This section describes the step-by-step time series analysis for counterfactual prediction of NO_2_ in California to illustrate the details of our approach.

**Figure S1.**
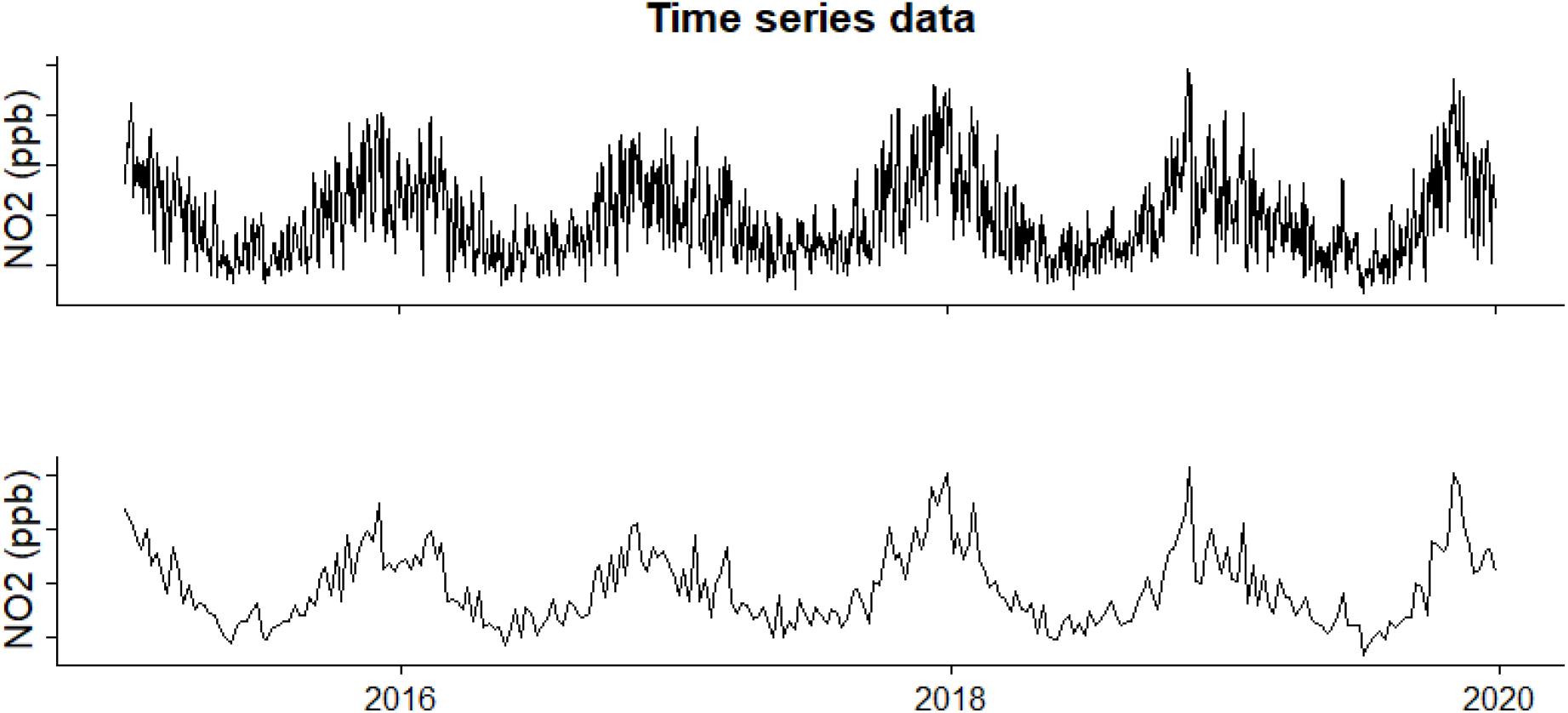
(a) Time series data for daily average NO_2_ levels for California for a five-year period. (b) Weekly averages of data in (a) used for training the SARIMA model.

Figure S1 (a) shows raw data for daily average NO_2_ levels for five years (January 1, 2015 to December 12, 2019). For our model, we performed weekly averages resulting in 261 data points for the five-year period, one time point for each week (Figure S1 (b)).

First, we created 1,000 bootstrapped time series from the weekly averaged training data using Box-Cox and Loess based decomposition.(33) Figure S2 shows a plot of 95% CI of these bootstraps.

**Figure S2.**
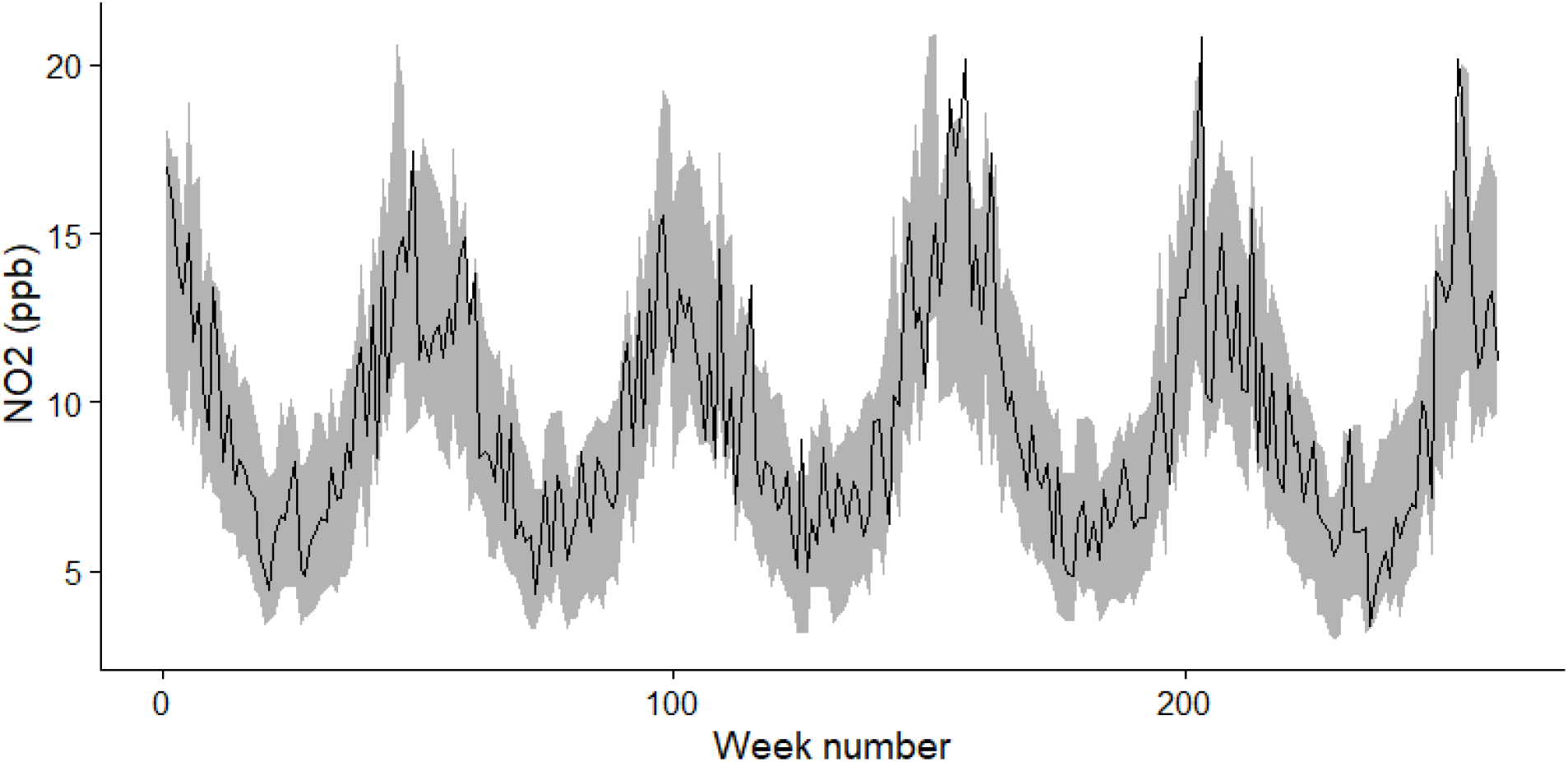
Time series data of weekly averages of NO_2_ levels in California (black line) with the confidence intervals of the 1,000 generated bootstraps (shaded area).

Next, we fit a SARIMA model to each of these bootstrapped time series, adjusting for meteorological factors, namely temperature, precipitation, and humidity. The average of the fitted values of the model is shown in Figure S3 along with 95% CI. We found that the model fits the data well with a mean absolute scaled error (MASE) = 0.8. Subsequently, we used the 1,000 fitted models to make counterfactual forecasts of weekly averages for 16 weeks in 2020 (January to April). Figure S4 shows the average predictions from the 1,000 models and the corresponding 95% CI. A comparison of the forecasts of the observed data shows that the NO_2_ levels begin to deviate from the predictions at about 10 weeks.

**Figure S3.**
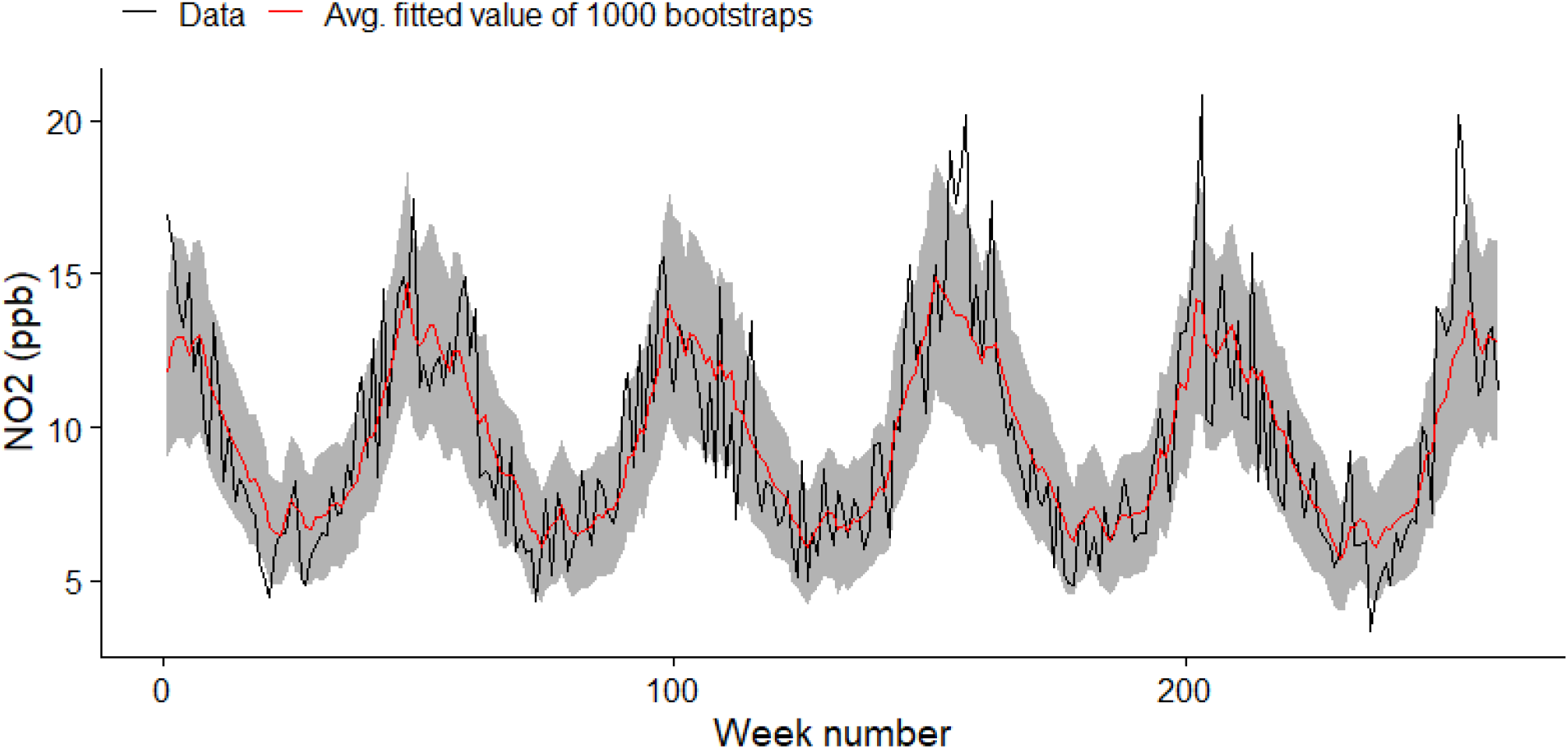
Average of the fitted values from 1,000 SARIMA models (red), corresponding95% CI (shaded area), and observed data (black).

**Figure S4.**
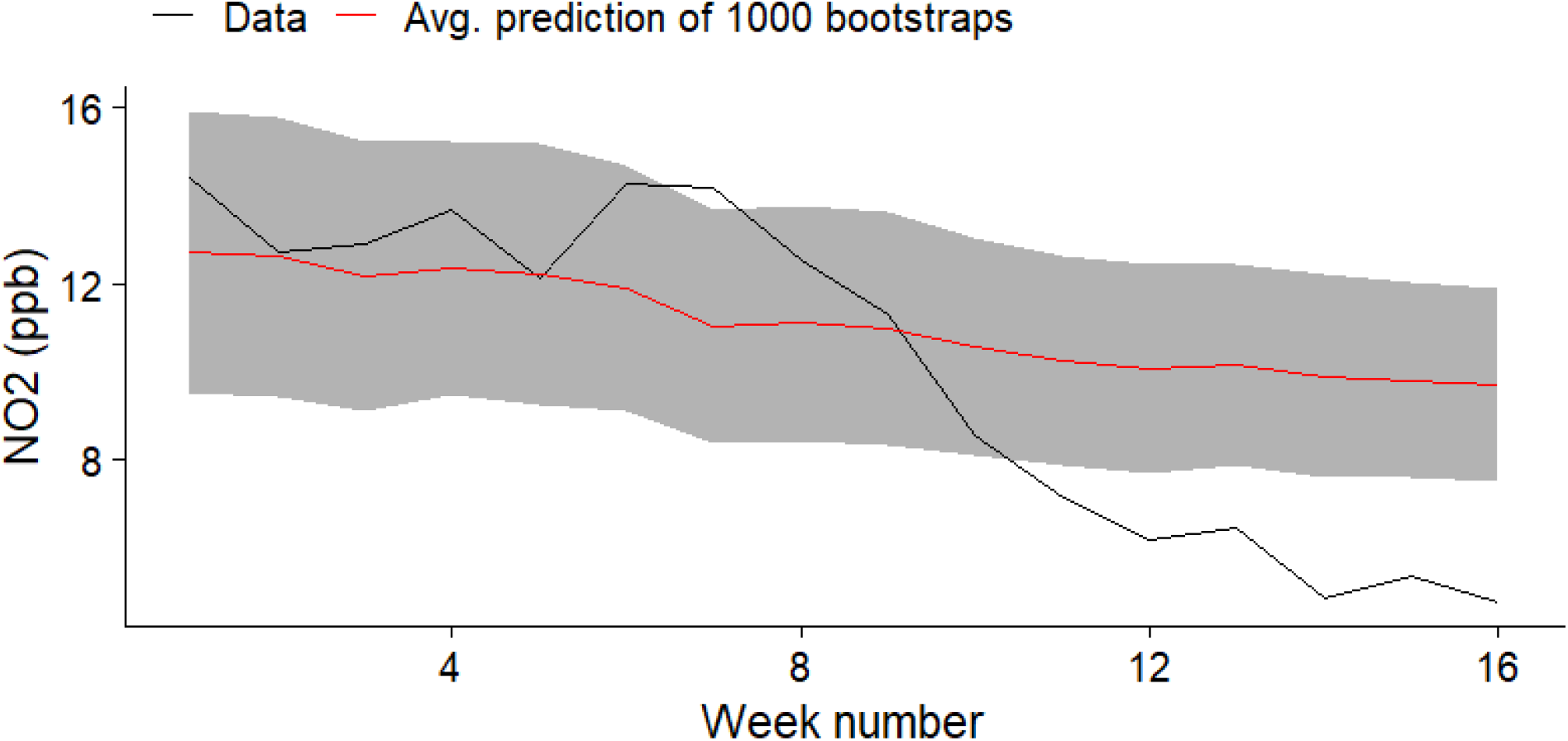
Average of predictions from 1,000 SARIMA models for 16 weeks in 2020 (red), corresponding 95% CI (shaded area), and observed data (black).

Finally, we calculated the difference between the average of the fitted predictions and the observed data (Figure S5). We found that the NO_2_ levels start deviating from our forecasts at the same time as the state of emergency was declared in California. The analysis described above was performed for each state and for PM_2.5_ and NO_2_.

**Figure S5.**
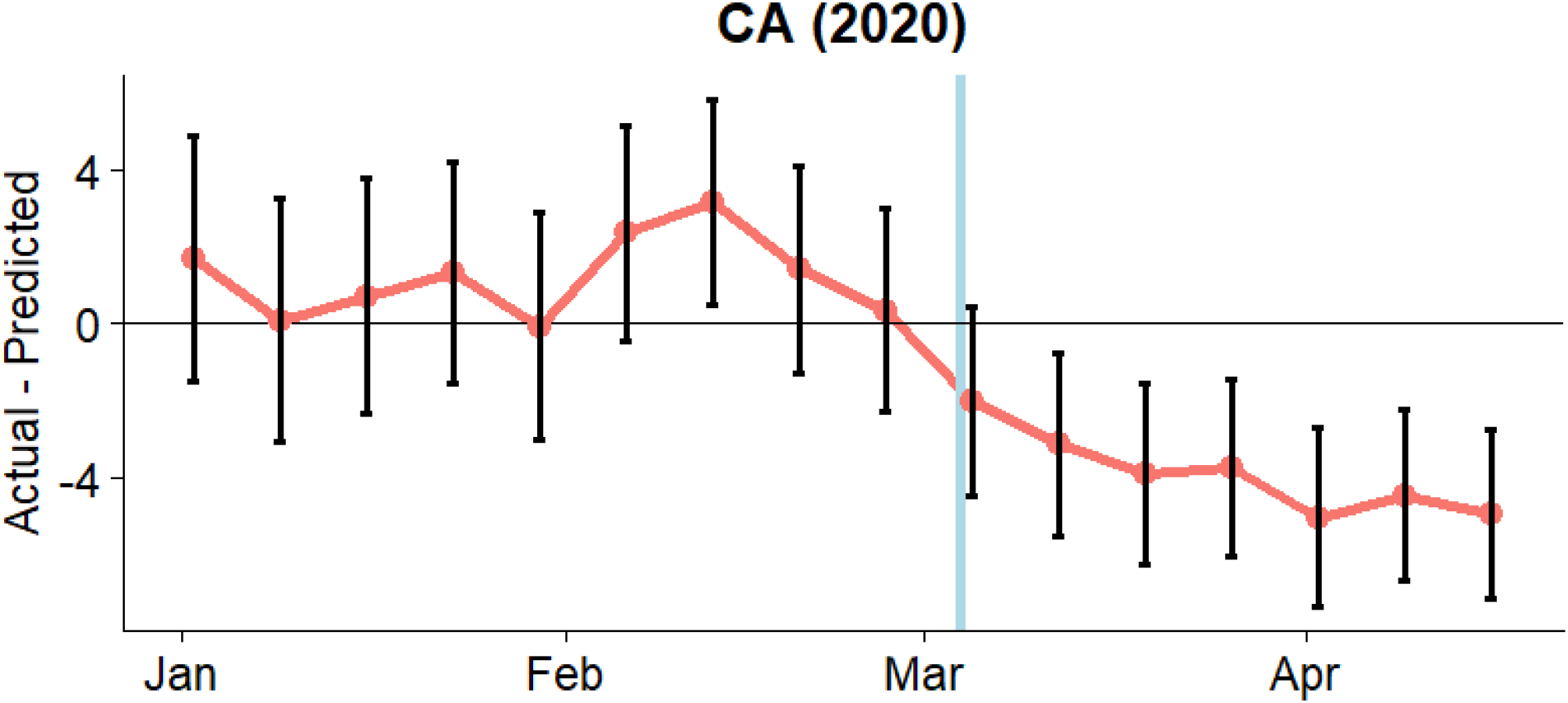
The weekly deviations between actual and predicted NO_2_ levels (ppb) for 16 weeks in 2020. The blue line corresponds to the day of declaration of a state of emergency in California.

## Model Evaluation

Figure S6 shows the MASE distribution for all fitted models.

**Figure S6.**
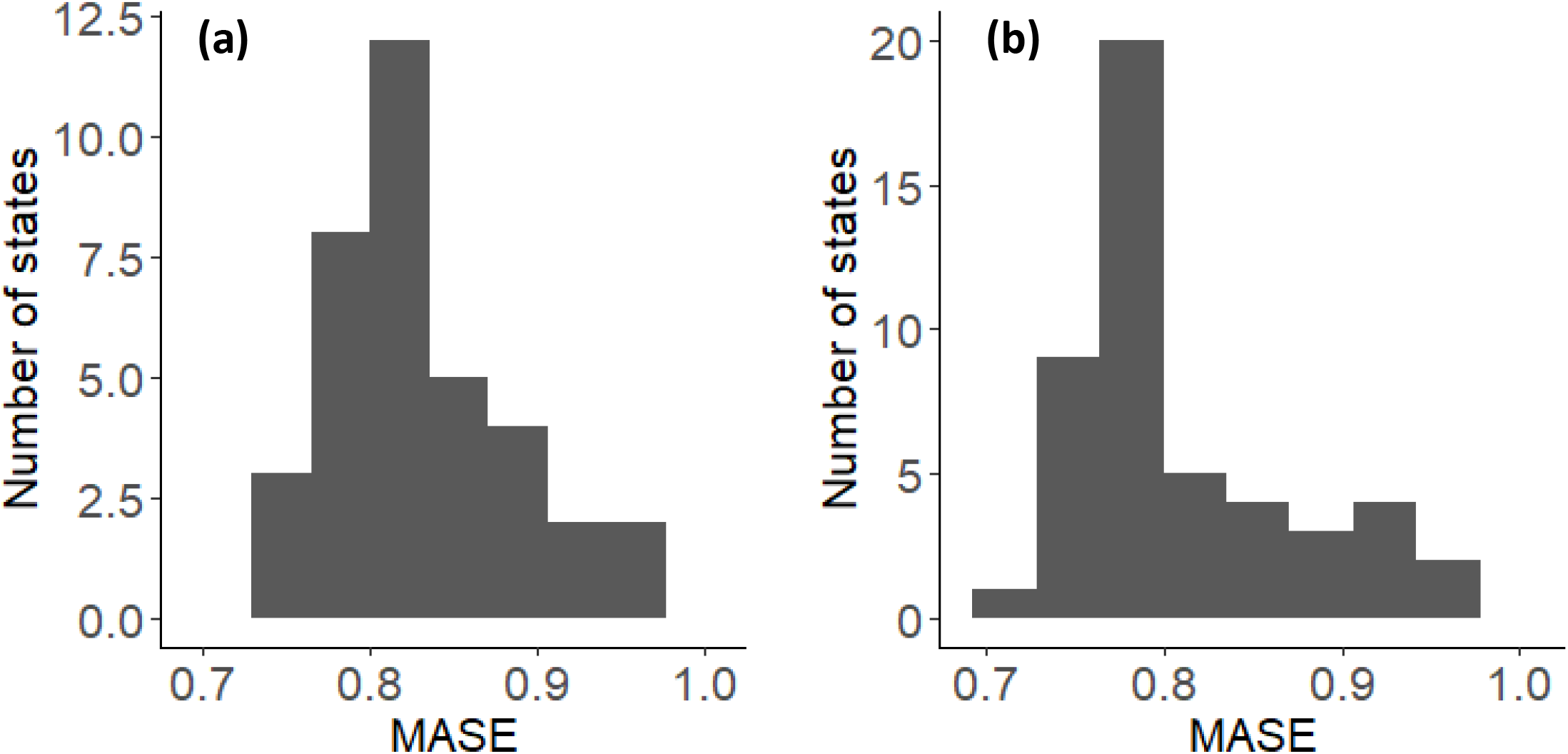
Mean absolute scaled error (MASE) for fitted models for all states for (a) NO_2_ and (b) PM_2.5_.

## National level pollutant emissions

**Table S2.** Annual PM_2.5_ and NO_2_ emissions (in tons) by emission source in the continental United States in 2014.(29)

| Sources | Emission (Tons) |  |
| --- | --- | --- |
|  | NO<sub>2</sub> | PM<sub>2.5</sub> |
| **Stationary** | 4,649,676 (34.8%) | 3,292,221 (61.4%) |
| **Mobile** | 7,513,708 (56.2%) | 336,554 (6.3%) |
| **Fire** | 291,213 (2.2%) | 1,730,360 (32.3%) |
| **Biogenics** | 903,124 (6.8%) | 0 (0.0%) |

## Tests for statistical significance in decrease in pollutant levels

**Table S3.** P-values obtained from Mann-Whitney test comparing the deviation from predicted values before versus after the declaration of a state of emergence in each state.

| State | P-value | State | P-value |
| --- | --- | --- | --- |
| AZ | < 0.001 | KY | 0.005 |
| CA | < 0.001 | MO | 0.007 |
| CT | < 0.001 | OK | 0.007 |
| MA | < 0.001 | WI | 0.008 |
| NV | < 0.001 | IN | 0.011 |
| NM | < 0.001 | IA | 0.011 |
| PA | < 0.001 | SC | 0.019 |
| RI | < 0.001 | MI | 0.021 |
| UT | < 0.001 | OR | 0.021 |
| VT | < 0.001 | KS | 0.028 |
| VA | < 0.001 | OH | 0.028 |
| NJ | 0.001 | MN | 0.057 |
| MS | 0.002 | WA | 0.071 |
| NC | 0.002 | CO | 0.074 |
| ND | 0.002 | GA | 0.09 |
| ID | 0.003 | WY | 0.09 |
| ME | 0.004 | MT | 0.356 |
| MD | 0.004 | FL | 0.604 |

**Figure S7.**
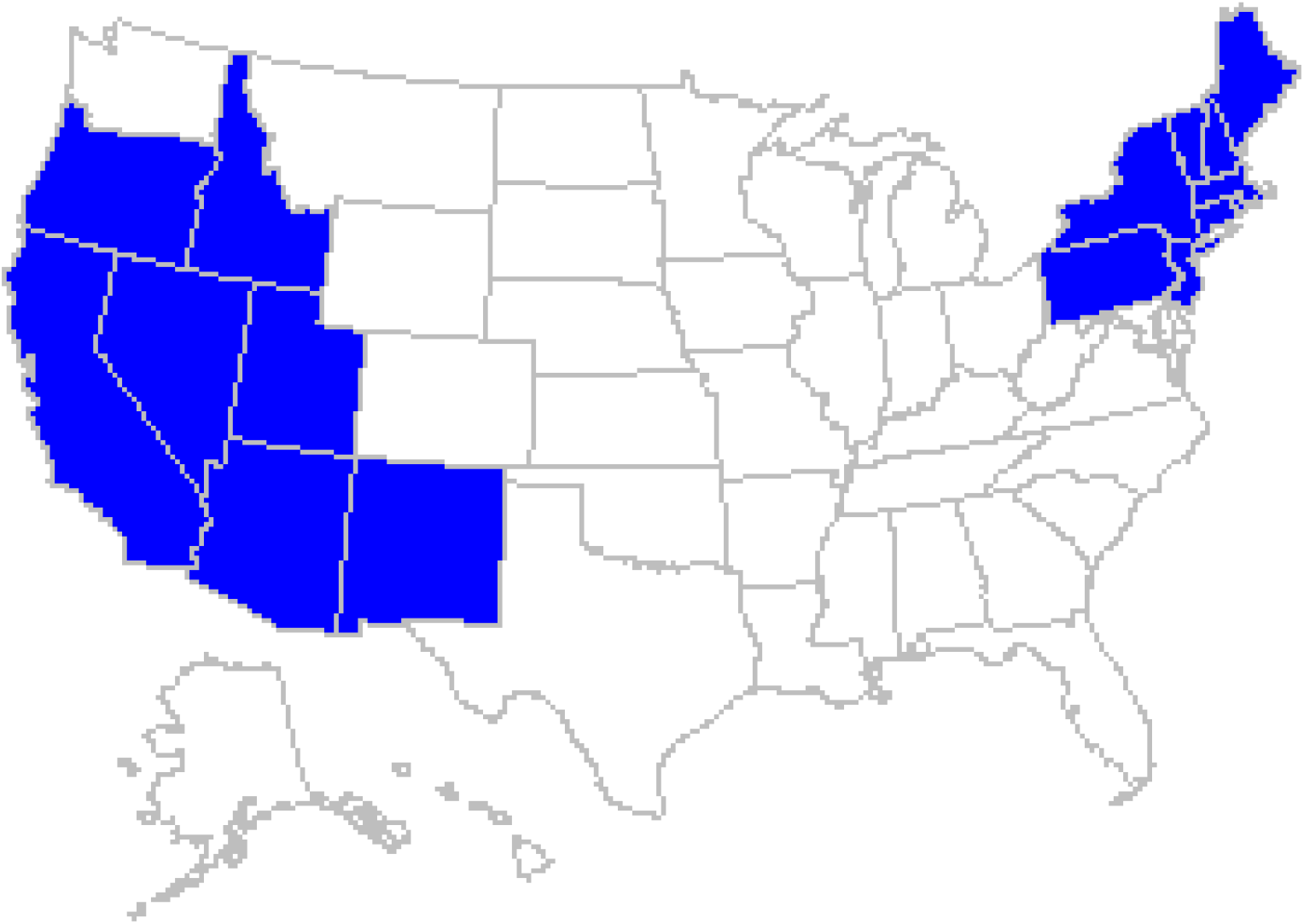
States with a statistically significant difference (P < 0.05) in deviations from predicted values before versus after the declaration of a state of emergency are highlighted in blue.

**Figure S8.**
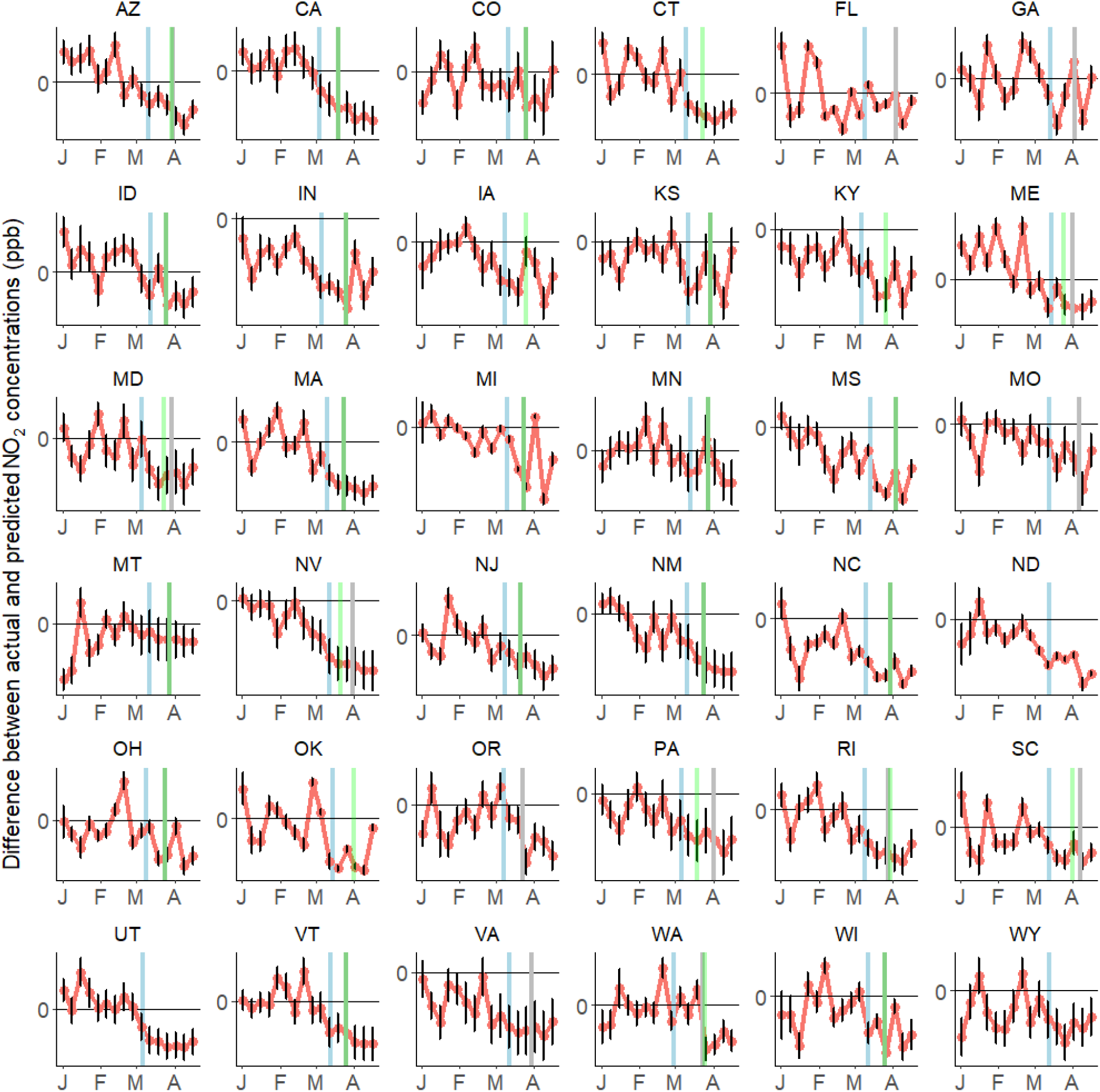
Weekly deviations between observed NO_2_ concentrations and counterfactual predictions (e.g., absent the pandemic) for each state. The predictions were made for 16 weeks from January 1 to April 23, 2020. The blue, green, and grey vertical lines mark the dates corresponding to the declaration of a state of emergency, non-essential business closures, and shelter-in-place/stay-at-home orders in each state.

**Figure S9.**
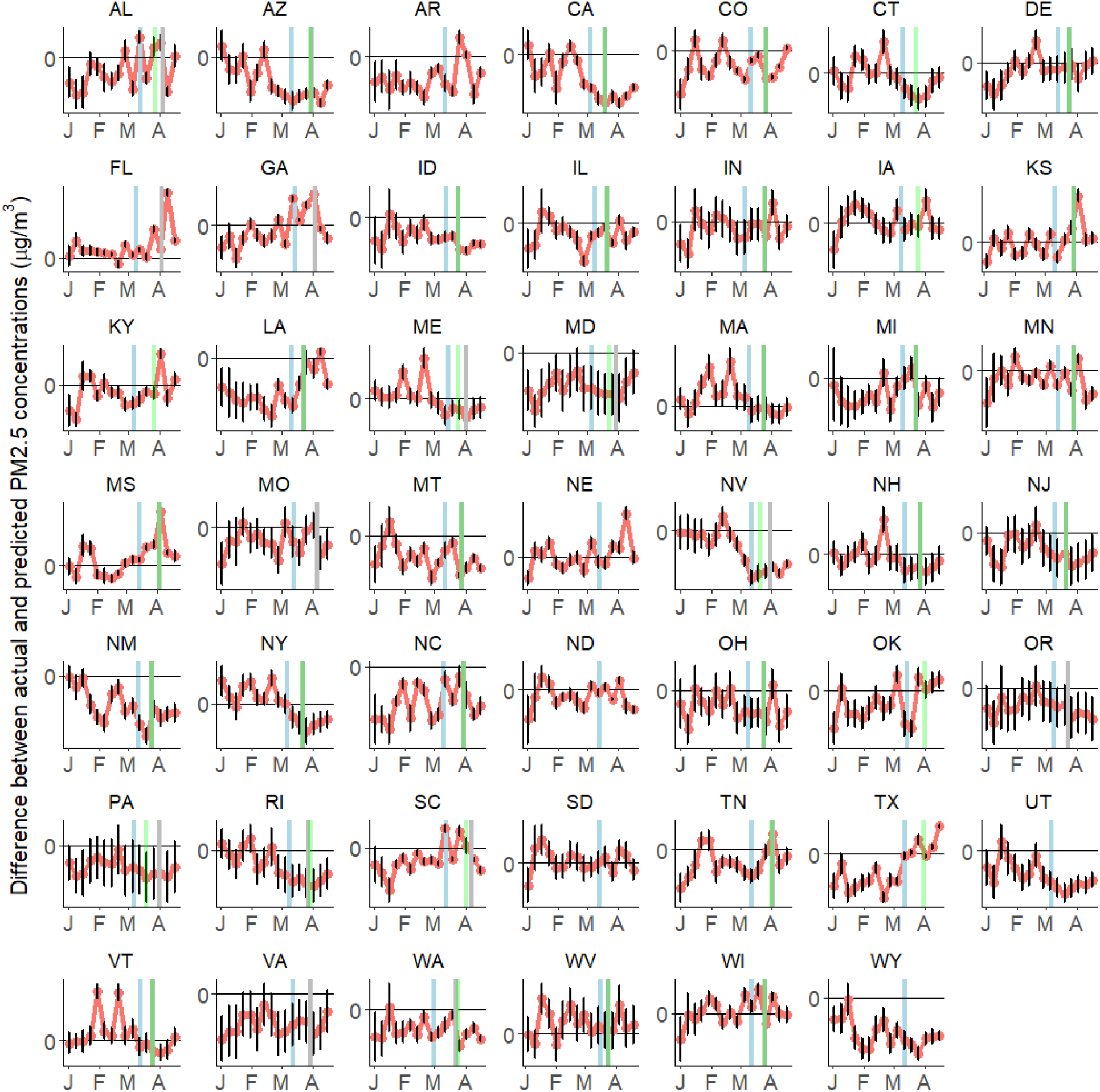
Weekly deviations between observed PM_2.5_ concentrations and counterfactual predictions (e.g., absent the pandemic) for each state. The predictions were made for 16 weeks from January 1 to April 23, 2020. The blue, green, and grey vertical lines mark the dates corresponding to the declaration of a state of emergency, non-essential business closures, and shelter-in-place/stay-at-home orders in each state.

